# Deep brain electrical neurofeedback allows Parkinson patients to control pathological oscillations and quicken movements

**DOI:** 10.1101/2020.06.10.20127829

**Authors:** Oliver Bichsel, Lennart H. Stieglitz, Markus F. Oertel, Christian R. Baumann, Roger Gassert, Lukas L. Imbach

## Abstract

Parkinsonian motor symptoms are linked to pathologically increased beta-oscillations in the basal ganglia. While pharmacological treatment and deep brain stimulation (DBS) reduce these pathological oscillations concomitantly with improving motor performance, we set out to explore neurofeedback as an *endogenous* modulatory method. We implemented *deep brain electrical neurofeedback* to provide real-time visual neurofeedback of pathological subthalamic oscillations measured through implanted DBS electrodes. All 8 patients volitionally controlled ongoing beta-oscillatory activity within minutes of training. During a single one-hour training session, the reduction of beta-oscillatory activity became gradually stronger and accelerated hand movements. Lastly, endogenous control over deep brain activity was possible even after removing visual neurofeedback, suggesting that neurofeedback-acquired strategies were retained in the short-term. We observed a similar motor improvement when the learnt mental strategies were applied 2 days later. Further improvement of deep brain neurofeedback might benefit Parkinson patients by improving symptom control, even in the absence of real-time neurofeedback.

## Introduction

Patients with Parkinson’s disease (PD) suffer from progressive impairment of motor function caused by a loss of dopaminergic neurones in the basal ganglia and brain-stem ^1^. Symptomatic treatments of PD include dopaminergic drugs or deep brain stimulation (DBS). In DBS, electrodes are surgically implanted into basal ganglia nuclei – typically the subthalamic nucleus (STN) or the globus pallidus internus (GPi) – both of which are relevant structures in affected movement modulatory circuits ^2^. Besides greatly and almost immediately alleviating motor symptoms through the application of high frequency stimulation, implanted DBS electrodes have also proved to be invaluable in the quest for better understanding PD, namely by shedding light on the electrophysiological hallmarks of PD through recording of deep brain local field potentials (LFP).

Characteristically, patients with PD show pathologically increased levels of beta-oscillatory activity ^3–5^. Beta-oscillations have been correlated with clinical measures of PD, with a reduction of signal power in the beta-band corresponding to a clinical improvement of motor symptoms ^6–8^. Furthermore, the two main therapeutic strategies, the administration of L-Dopa ^3,9^ and high-frequency DBS ^8,10^, both lead to a suppression of beta-synchronicity in the STN. Beta-oscillations show fast and movement-dependent modulation over time ^11,12^ and can serve as a biomarker and feedback signal to control the delivery of DBS ^13^.

Besides the *exogenous* manipulation of subthalamic oscillatory activity through pharmacotherapy and DBS, it remains to be unveiled whether *endogenous* modulation is also achievable. Neurofeedback has emerged as a promising, *endogenous* technique enabling self-regulation of ongoing brain activity. It relies on the real-time extraction of relevant features from neuronal activity, that is presented to the subject. When applied to deep brain structures, neurofeedback might enable the control of brain activity in areas that fall outside the scope of conscious awareness. In other words, through mirroring ongoing neuronal deep brain activity, neurofeedback empowers people to explicitly modify certain brain states. Such a real-time self-modulation would be especially useful in sudden exacerbations of symptoms, which are characteristic for PD.

The modulation of deep brain activity through neurofeedback is not a new notion and has already been explored by a large body of real-time functional magnetic resonance imaging (rtfMRI) research, providing for instance evidence for voluntary control over the substantia nigra/ventral tegmental area complex (SN/VTA) through mental imagery ^14^. However, compared to LFP recordings, rtfMRI is confined to the laboratory environment, suffers from a time lag caused by the haemodynamic response and is limited by a low temporal resolution. Using LFP recordings from DBS electrodes, a recent study showed that the 5-minutes average resting subthalamic beta-activity after 10 minutes of neurofeedback differed with respect to the resting period before neurofeedback^15^, suggesting short- to mid-term effects of neurofeedback. Nevertheless, it remains to be unveiled whether deep brain electrode-guided neurofeedback allows for real-time modulation of ongoing subcortical activity as well as whether modulatory effects improve with exposure time, are maintained after removal of neurofeedback and result in a better motor outcome.

We hypothesise that by providing short-delay, electrical deep brain neurofeedback, PD patients can learn to volitionally control the amount of pathological subthalamic beta-oscillations in *real-time* resulting in a better motor performance. Ultimately, *endogenously* modulating pathological deep brain oscillations could serve as a new approach to reduce Parkinsonian motor symptoms.

## Materials and Methods

### Study cohort and surgery

We included ten PD patients (Table 1) undergoing bilateral DBS lead (Model 3389, Medtronic Neurological Division, Minneapolis, MN, USA) implantation into the STN. DBS leads were implanted after MRI-based direct targeting of the STN. Accurate implantation within the STN was intraoperatively verified by micro-electrode recordings (Leadpoint, Medtronic Neurological Division, Minneapolis, MN, USA) in steps of 0.5 mm until target (starting 10 mm above the target), clinical response upon intraoperative stimulation and intraoperative computed tomography (CT). We verified accurate electrode placement by comparing the planned and actual electrode position in the postoperative CT scan. All experiments were performed on the second postoperative day in L-Dopa OFF and DBS OFF state, prior to implantation and connection of the stimulation decive (Activa PC, Medtronic Neurological Division, Minneapolis, MN, USA) 3–5 days after electrode placement. Decisions on patient selection and surgical procedures were not affected by this study and exclusively based on clinical grounds. This study was approved by the local ethics committee (Kantonale Ethikkommission Zürich, board decision on the amendment to KEK-ZH: 2012– 0327), and all patients gave informed written consent prior to inclusion.

**Table 1:** Demographics and clinical patient characteristics. MDS-UPDRS: Movement Disorder Society-Unified Parkinson’s Disease Rating Scale; ON/OFF values of preoperative L-Dopa challenge test; LED: preoperative levodopa equivalent dose ^16^. The third part of the MDS-UPDRS includes the clinician-scored, monitored motor evaluation (33 scores based on 18 items) ^17^, ranging from the worst MDS-UPDRS III score of 132 (= 4·33) points to 0 points, with all items scoring the same as in healthy individuals. The minimal clinically important difference on the UPDRS motor score is 2.3–2.7 points ^18^.

| ID | Age [y] | Gender | PD type | Side dominance | Disease duration [y] | Hoehn-Yahr Scale | MDS-UPDRS III ON/OFF | LED [$\frac{\text{mg}}{\text{d}}$] |
| --- | --- | --- | --- | --- | --- | --- | --- | --- |
| 1 | 70 | M | equivalent | left | 10 | 2.5 | 37/62 | 2'340 |
| 2 | 63 | M | akinetic-rigid | right | 7 | 2.5 | 27/44 | 1'800 |
| 3 | 45 | M | tremor | right | 2 | 1.5 | 19/28 | 375 |
| 4 | 60 | M | tremor | right | 19 | 1.5 | 17/41 | 1'750 |
| 5 | 62 | F | akinetic-rigid | right | 10 | 2.5 | 29/46 | 275 |
| 6 | 65 | M | equivalent | right | 9 | 2 | 20/32 | 1'240 |
| 7 | 57 | F | equivalent | right | 9 | 2 | 25/54 | 620 |
| 8 | 71 | M | tremor | left | 4 | 2.5 | 19/35 | 1'350 |
| 9 | 58 | F | equivalent | right | 13 | 2.5 | 20/33 | 1'500 |
| 10 | 48 | M | equivalent | left | 5 | 2 | 24/43 | 1'525 |

### Experimental setup

We measured STN LFPs from temporarily externalised DBS wires (4 electrode sites per lead) sampled at 5 kHz by a BrainAmp DC (Brain Products GmbH, Gilching, Germany) EEG recorder. A ground and a reference scalp EEG input were placed over the central midline region C_z_ (according to the international 10/20 system) and connected to the EEG recorder. Signal quality was checked visually for electrode or movement artefacts. All experiments were performed in a sitting or supine position such that patients were able to see the computer screen providing visual feedback. As a baseline condition, we first recorded a resting state condition in which patients looked at the blacked monitor for 60 s. Using the BrainVision Recorder and Analyzer (both from BrainProducts GmbH, Gilching, Germany), we then identified the pair of adjacent electrode contacts contralateral to the PD-dominant hand that showed the highest beta power in the power spectral density – similar to ^11^ – and determined the beta-peak frequency as an individual parameter to be used for signal processing.

### Visual neurofeedback

We implemented a custom-written C++-programme to extract an instantaneous beta-power estimate from the externalised leads. The programme built on the BrainAmp Software Development Kit (Version 002, BrainProducts GmbH, Gilching, Germany), which provided a framework allowing direct access to the BrainAmp EEG amplifier through the USB port. Every 40 ms, 200 samples (0.04 s * 5 kHz) per channel were bussed through the USB port for processing of the raw signal: First, a bipolar channel was created by subtracting one channel from its adjacent channel; second, the raw bipolar lead signal was band-pass filtered (Butterworth, 3^rd^ order) around the previously determined individual beta peak-frequency (5 Hz) by a rational transfer function; third, the signal was rectified; fourth, the signal from this and the previous 4 blocks was averaged (running window average). Finally, the resulting value, representing the instantaneous smoothed beta-activity over the preceding 200 ms, was updated at a rate of 25 Hz and used for visual neurofeedback. This real-time neuro-feedback parameter determined the location of a disc that moved horizontally from lower values on the left of the screen to higher values on the right (Fig. 1). The lower limit of the screen was defined as the 75^th^ percentile beta-power value obtained during 1 minute of overt fist opening and closing with the PD-dominant hand. The upper limit of the screen was defined as the 75^th^ percentile neurofeedback value during 1 minute of staring at the blacked computer screen.

**Figure 1:**
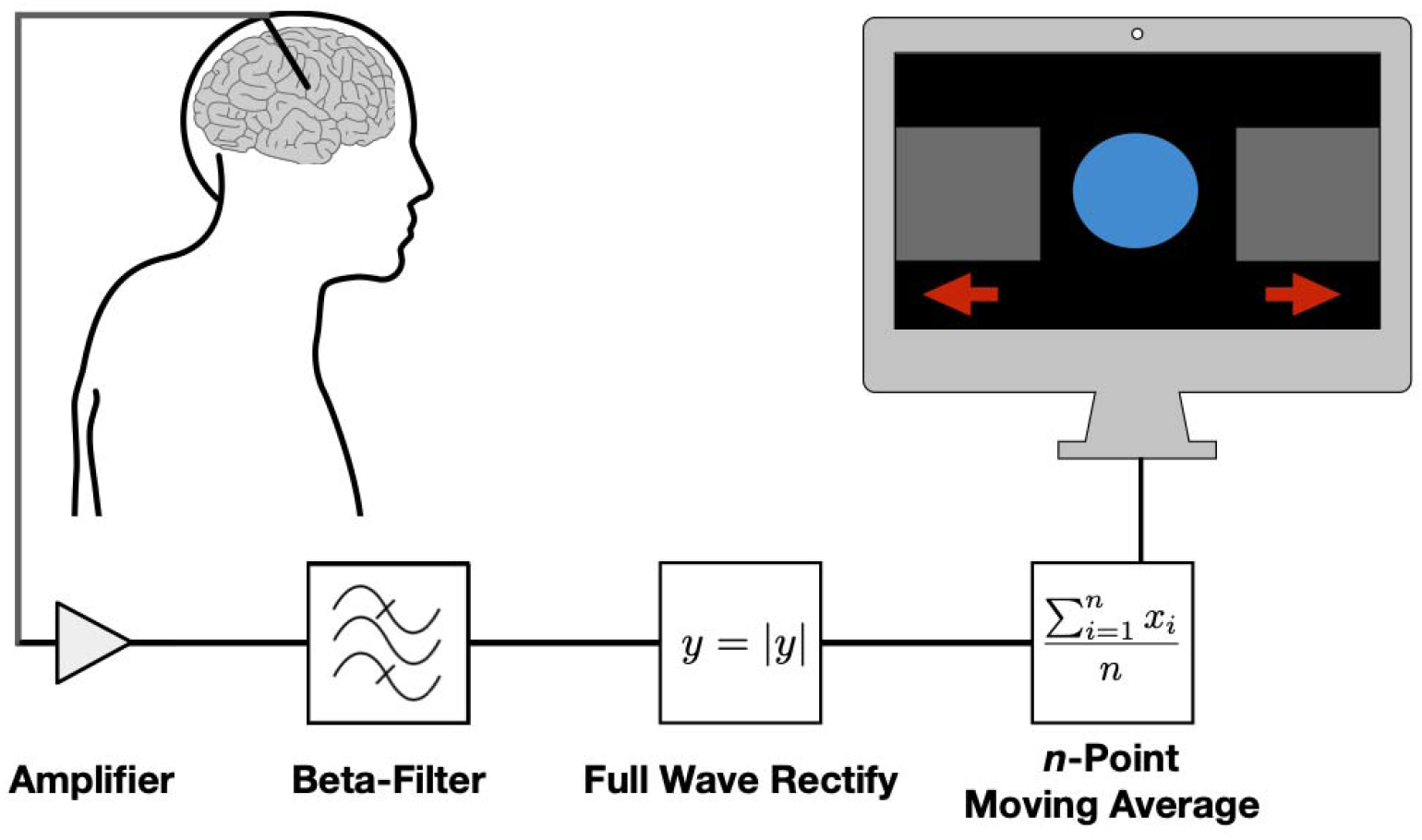
Deep brain electrical neurofeedback loop. Deep brain stimulation electrodes (upper left corner) implanted into the subthalamic nucleus allowed for brain oscillation measurements. The raw, analogue signal was amplified and then digitally band-pass filtered around the individual beta-peak (± 5 Hz), rectified, averaged and used to control the position of the blue disc on the monitor (upper right corner). Patients tried to volitionally control the position of the disc, thereby closing the neuro-feedback loop while controlling pathological beta-activity.

### Neurofeedback experiment

The neurofeedback experiment was divided into three main parts, each comprising several 1-minute blocks: the pre-neurofeedback, the neurofeedback and the transfer part (Supplementary Figure 1). The pre-neurofeedback part consisted of one block of baseline (staring at the blacked computer screen), followed by a block of downregulation, where patients were asked to think about fluent movements (motor imagery); followed by a block of upregulation, where patients were asked to think about bradykinetic movements or abruptly ending movements; followed by a block of baseline, upregulation and finally downregulation. For the neurofeedback (NF) rounds, we provided visual neurofeedback for all blocks except for the baseline blocks. During neurofeedback, patients were instructed to voluntarily move the disc on the screen to the right side of the screen (upregulation) or to the left of the screen (downregulation) without overt movements (visually controlled by the examiner). The study was single-blinded, i. e. patients were unaware of the neuromodulatory effect (beta-upregulation vs. downregulation) of moving the disc to either side. Patients were allowed to adapt their mental strategies depending on success. The following sequence of blocks was repeated three times (NF1, NF2 and NF3; except for Patient 1, in which NF2 had to be omitted due to time restrictions): baseline, downregulation, upregulation, baseline, upregulation, downregulation. In total, each patient underwent 30 blocks (24 blocks for patient 1) resulting in a raw experimental time of 30 minutes (24 minutes for patient 1). Allowing for short transition periods between the blocks, the total experimental time was usually between 40 and 50 minutes. Moreover, we noted the final mental strategies used by the patients to either up- or downregulate during the last round of neurofeedback (NF3). Since the bidirectional neurofeedback design allowed for intra-individual controls, differences in the amount of beta-oscillations would be solely attributable to the neurofeedback condition, making a separate control group superfluous. Moreover, the bidirectional neurofeedback design also controlled for possible eye movement contributions. For the transfer round, the computer screen was again turned off (no neurofeedback) and patients were instructed to reuse the previously learnt mental strategies for either up- or downregulating beta-activity while staring at the black computer screen and avoiding movements. The sequence during the transfer round was: baseline, downregulation, upregulation, baseline, upregulation, downregulation. To capture the behavioural effects of neurofeedback, each block during NF3 was immediately followed by a period of 15 s, where patients were instructed to alternatingly and completely (i. e. 180 ° rotations) pro- and supinate their more severely affected hand as fast as possible. We used inertial measurement units (IMUs) mounted on the hand to log the recordings at 200 Hz (ZurichMOVE, www.zurichmove.com). Two days after the initial neurofeedback experiment (i. e. after DBS lead internalisation and thus without neurofeedback), we assessed the effect of neurofeedback-learnt mental strategies on motor performance (again in L-DOPA OFF and DBS OFF). The experimental sequence for this long-term transfer experiment was the same as in NF3, but without visual neurofeedback: baseline, downregulation, upregulation, baseline, upregulation, downregulation, with the pro- and supination motor task following each block.

### Post-hoc data analysis and statistics

The raw signal from each electrode site was saved in the BrainAmp EEG format and later converted to European Data Format (edf) by BrainVision Analyzer (BrainProducts GmbH, Gilching, Germany). The edf was imported into MATLAB (R2018b, The MathWorks Inc., Natick, Massachusetts, USA) for all further data analyses and statistics. To reconstruct the previously visualised signals, we calculated the same bipolar montage from the raw data and filtered using the same algorithms as during the real-time programme. The signal from each block was later rectified and averaged.

The averaged values obtained for the up- or downregulation blocks were then normalised with the values from the preceding baseline block. Finally, the normalised values from the same condition of the same block (pre-neurofeedback, NF1, NF2, NF3, transfer) were averaged for between condition comparison. Hypothesis testing with two-sided Student’s *t*-test was used to determine whether neurofeedback significantly modulated the instantaneous beta-power. We used Wilcoxon’s signed rank test to compare the pre-neurofeedback with the transfer values. We correlated the MDS-UPDRS III OFF score with the beta-modulatory capacity to assess the relationship between disease burden and the ability to modulate pathological oscillations. The gyroscope data from the IMUs was imported into MATLAB. A custom-written script summed the three pairwise-orthogonal gyroscope axes up before integration and band-pass filtering between 0.25 and 4 Hz (3^rd^ order Butterworth). To examine the motor performance, the pro- and supination segments were visually identified for each patient and truncated to the first 12 s (informed decision to avoid the effect of prematurely terminated movements). The following metrics were calculated and averaged over repetitions of the same condition for each individual patient: number of pronosupination cycles (measure of frequency), mean peak angular amplitude (measure of movement extent), cumulative angular displacement (measure of total movement extent) and mean absolute angular acceleration (measure of the torque generated by the antagonistically acting pro- and supinator muscles). We used Wilcoxon’s signed rank test to compare the behavioural metrics after rest versus downregulation.

## Results

From the 10 patients included in this study, we had to exclude two patients because of intermittent high amplitude artefacts in the raw DBS signal, resulting in a total number of 8 patients included for data analysis.

### Beta-activity estimate during downregulation vs. rest

We calculated average beta-activity estimates normalised by the beta-activity estimate during the preceding rest block for each individual patient during downregulation in the baseline condition (pre-NF), all active neurofeedback rounds (NF1, NF2, NF3) and the transfer round (Fig. 2). The sample mean is below 1 for all but the pre-NF round, indicating that the beta-activity estimate was reduced compared to baseline. There is a significant reduction (Student’s *t*-test) for NF2 (*p* = 0.0294), NF3 (*p* = 0.00092) and the transfer round (*p* = 0.0134). The beta-activity estimates for the transfer round were also significantly lower than during the pre-NF round (Wilcoxon’s signed rank test, *p* = 0.0156). The variance decreases from NF1 up to NF3 and again increases for the transfer round. More pronounced beta-modulations were achieved by patients with heavier disease burden (Fig. 3, significant relationship: *p* = 0.0083; Pearson correlation coefficient: *R* = *0*.84).

**Figure 2:**
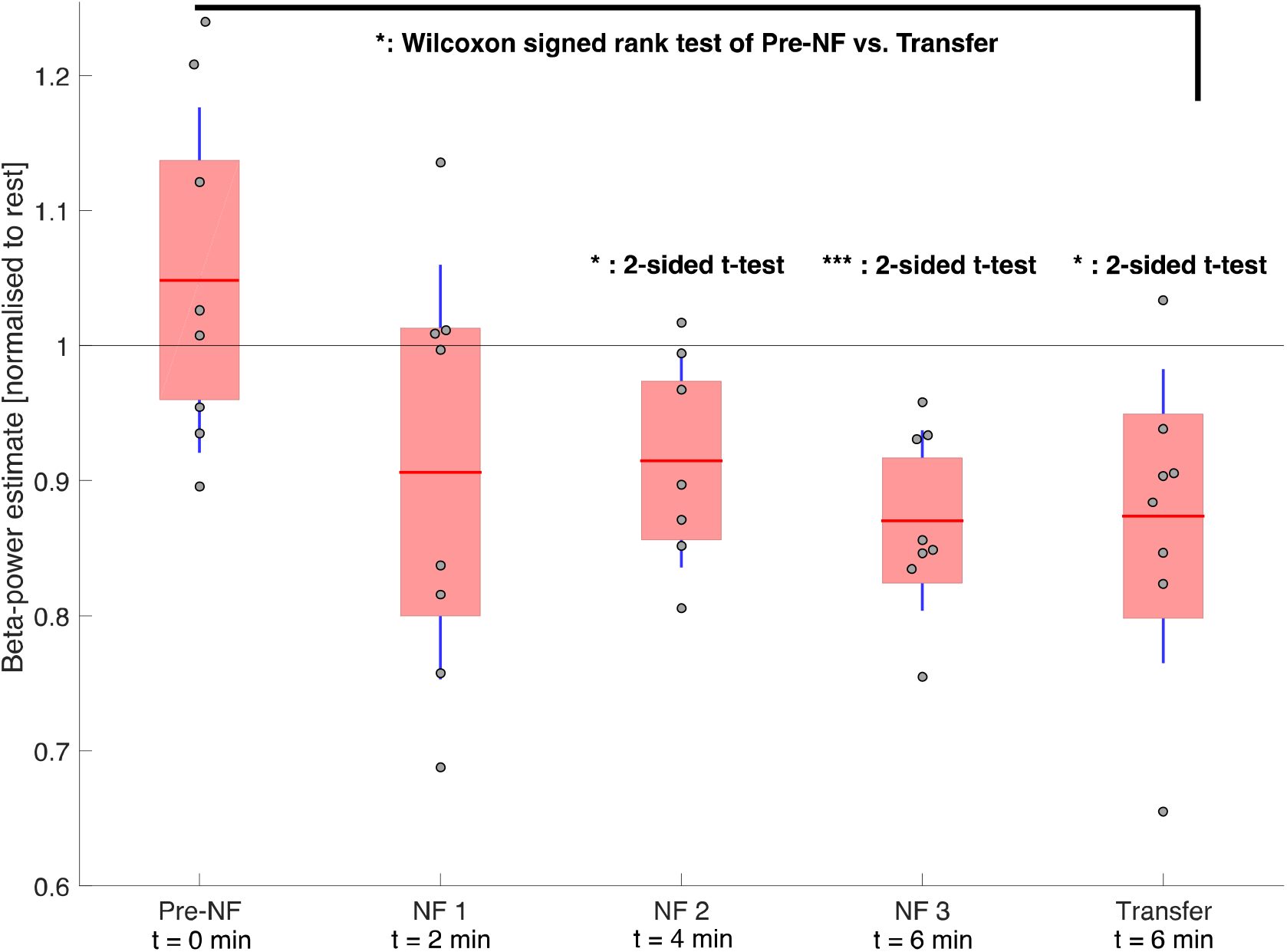
Learning to downregulate beta activity with DBS-neurofeedback. Beta-power estimates from all patients during downregulation – normalised to the beta-power estimate during each patients’ preceding rest block – are shown for the baseline (pre-NF), neurofeedback (NF1, NF2, NF3) and transfer rounds. We indicate the total amount of time *t* that patients have spent learning downregulation through neurofeedback until the end of that respective round. The group means are represented by the horizontal red lines, the standard deviations by the vertical blue lines and the 95 % confidence intervals by the red patched areas. We used the two-sided Student’s *t*-tests to test for significant beta-reductions compared to the baseline rest (horizontal black line at 1). Wilcoxon’s signed rank test was used to compare the dependent samples from the transfer round with their pre-NF value. *: p < 0.05; ***: p < 0.001.

**Figure 3:**
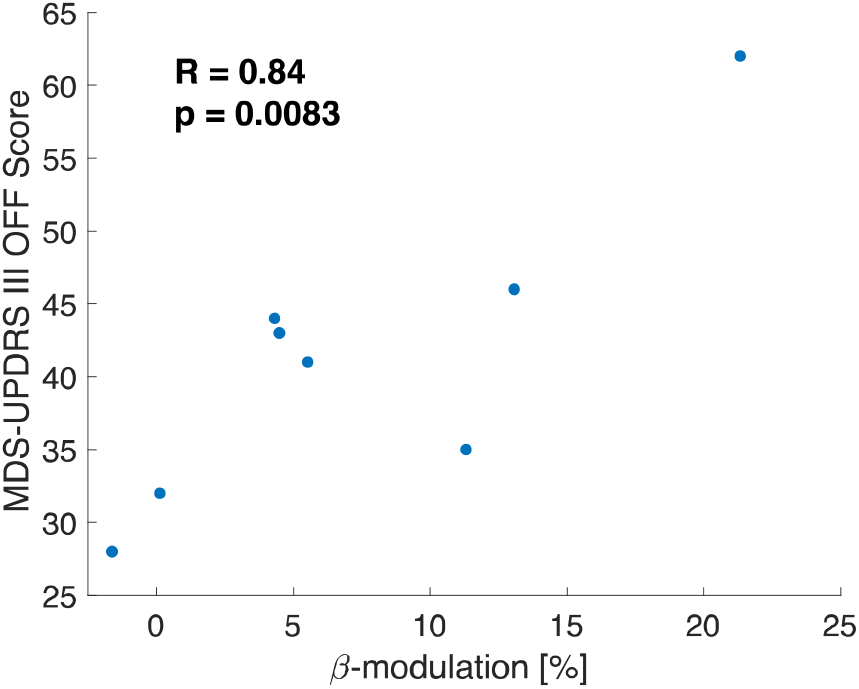
Relationship between disease burden and beta-modulatory capacity. We correlated the MDS-UPDRS III OFF score with the beta-modulatory capacity of each patient (*n* = 8) and found a significant relationship (*p* = 0.0083) with a positive Pearson correlation coefficient (*R* = 0.0083).

### Beta-activity estimate during upregulation vs. downregulation

The average beta-activity estimates during upregulation in the pre-NF, NF1, NF2, NF3 and the transfer round were calculated and normalised by the beta-activity estimate during downregulation of the same round (Fig. 4). The sample means were above 1 for all neurofeedback rounds, meaning that the beta-activity estimates were increased as compared to downregulation. For NF3, the sample mean was significantly increased in Student’s *t*-testing (*p* = 0.0288).

**Figure 4:**
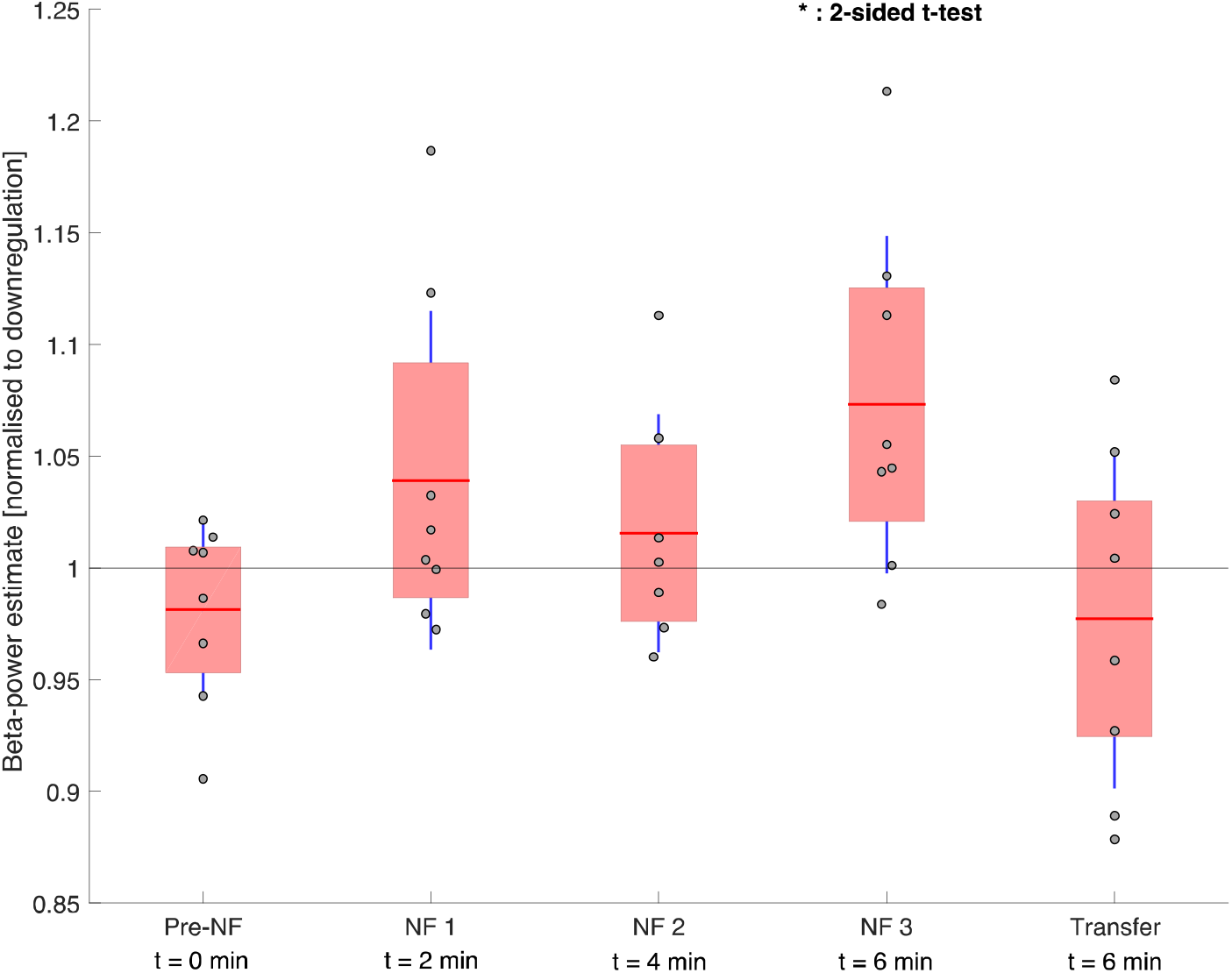
Learning bidirectional neurofeedback. Beta-power estimates from all patients during upregulation – normalised to the beta-power estimate during each patients’ downregulation condition – for the Pre-NF, NF1, NF2, NF3 and Transfer rounds show increasing beta activity upon neurofeedback aided upregulation. We indicate the total amount of time *t* that patients spent learning upregulation through neurofeedback until the end of that respective round. The means are represented by the horizontal red lines, the standard deviations by the vertical blue lines and the 95 % confidence intervals by the red patched areas. We used two-sided Student’s *t*-testing to determine significant beta-reductions compared to the downregulation condition (horizontal black line at 1). *: p < 0.05.

### Mental strategies used

The final mental strategies used for up- or downregulating beta-activity were all different from the initially suggested ones and highly personalised: Strategies for down-regulation ranged from imagining running, walking or cycling to just ‘moving the disc to the left’; Strategies for upregulation included imagining abrupt movements, difficulties in initiating a movement, freezing, freezing during catching a fish, relaxation and trying to ‘move the disc to the right’.

### Behavioural outcome after neurofeedback and long-term retention of mental strategy

To test for a behavioural effect of neurofeedback-learnt mental downregulation strategies, a set of movement metrics for the first 12 s of pronosupination during the rest condition was compared with the downregulation condition in 7 of the 8 patients (failure of recording in 1 patient). The movement frequency was significantly (Wilcoxon’s signed rank test, *p* = 0.0312) improved after downregulation (Fig. 5A left). To test for a bias towards smaller movement amplitudes, we calculated the mean peak angular amplitude, which was slightly and non-significantly reduced (Fig. 5B left).

**Figure 5:**
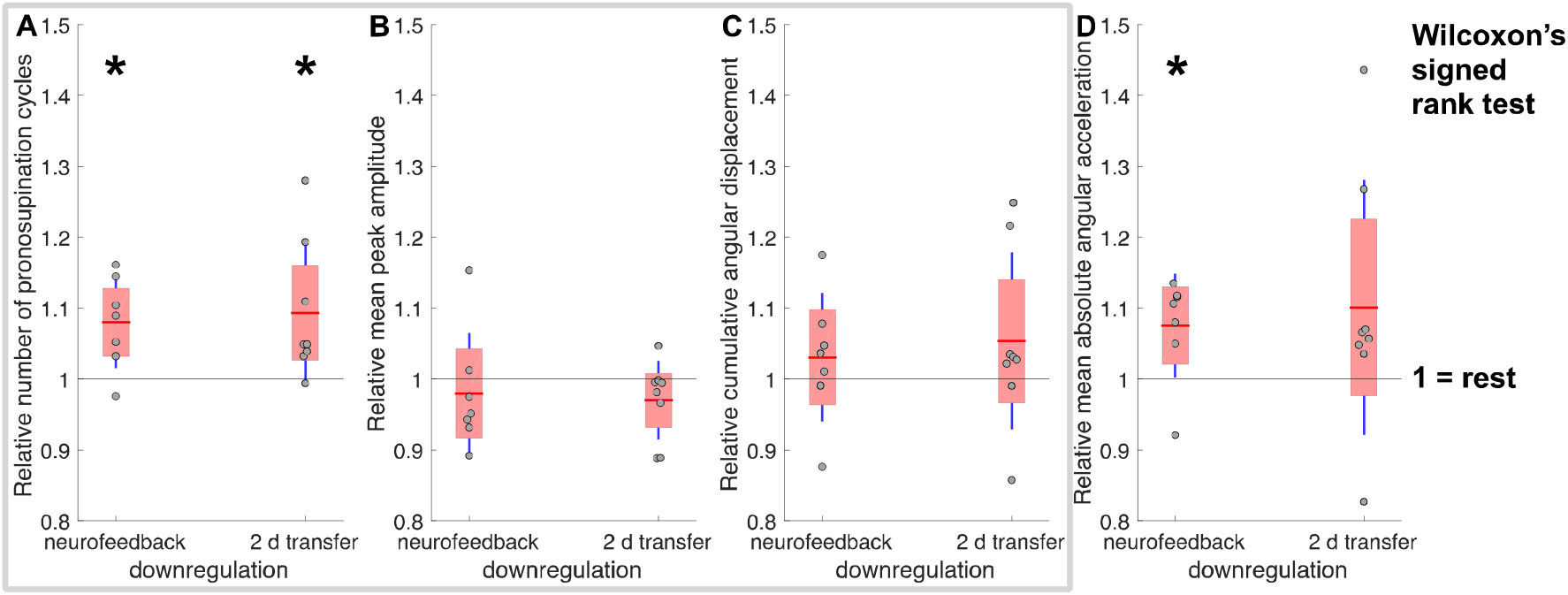
Behavioural output metrics during the first 12 s of pronosupination during NF3 (neurofeedback downregulation) and long-term (2 d) mental strategy transfer downregulation. (A) The pronosupination frequency (proportional to the number of cycles during the first 12 s of pronosupination) was significantly increased after downregulation as compared to rest (*p* = 0.0312 and *p* = 0.0156 during NF3 and 2 d mental strategy transfer, respectively) while (B) the mean peak angular amplitude was only slightly and non-significantly decreased. (C) The cumulative angular displacement (i. e. degrees travelled) was generally higher after downregulation as compared to rest. The grey rectangle highlights the mutual dependence between the three variables in (A), (B) and (C), i. e. frequency * mean peak angular amplitude = cumulative angular displacement. (D) The torque generated by the antagonistically acting pro- and supinator muscles was estimated through the mean absolute angular acceleration, which was significantly increased after downregulation as compared to rest during the neurofeedback experiment (*p* = 0.0469).

The cumulative angular displacement, a combination of the movement frequency and the mean peak angular amplitude, still favoured the downregulation condition (Fig. 5C left). Furthermore, the mean angular acceleration was significantly increased after downregulation as compared to the rest condition (Wilcoxon’s signed rank test, *p* = 0.0469), indicating a significantly higher mean torque generated after downregulation (Fig. 5D left). We also examined the motor performance after application of neurofeedback-learnt and retained mental strategies in all 8 patients after 2 days, when DBS leads had already been internalised, and found a similar pattern (Fig. 5; Wilcoxon’s signed rank test for the number of pronosupination cycles, *p* = 0.0156).

## Discussion

Many studies have provided evidence for a mutual interaction between beta-oscillatory activity in the STN and motor symptoms in PD. Inspired by real-time fMRI studies providing proof-of-principle for endogenous modulation of deep brain networks (although cumbersome and not transferable into everyday life), we investigated a novel neurofeedback technique using DBS electrodes implanted in the STN. Using this approach, we provided real-time direct visual neurofeedback of subcortical beta-power in the form of a disc that moved horizontally on a screen, thereby empowering patients to control their own pathological deep brain oscillations. The core findings of this study were that deep brain electrode-guided neurofeedback allows PD patients to gain control over subthalamic beta-oscillations, that this control is gradually improved with neurofeedback training and has a positive effect on motor performance. Furthermore, our findings suggest that neurofeedback-learnt strategies can be retained in the short and long-term as well as a beneficial effect on motor performance observed 2 days after neurofeedback-learning.

### Deep brain electrical neurofeedback quickly enables control over deep brain activity

Here, we demonstrate that real-time modulation of beta-activity is possible, even within as little as 4 minutes of downregulation training (*p* = 0.0294) and 6 minutes of upregulation training (*p* = 0.0288). This minimal-latency, endogenous access to deep brain activity greatly outperforms rtfMRI neurofeedback studies, where the control over the BOLD signal requires at least 30 minutes of training. In comparison to EEG neurofeedback studies, that require multiple sessions for controlling cortical activity^19^, DBS neurofeedback seems to provide faster and more reliable strategy development for the participants. This striking difference could be explained by the direct and local measurement of neuronal activity, providing a source signal that is closer to the most affected brain region (basal ganglia) prior to network modulation through basal ganglia-cortical loops ^11,20^. More so, electrical neurofeedback of the STN seems to be readily achievable, as all patients in our study managed to reduce pathological oscillations after only 6 minutes of downregulation training.

### Control over deep brain activity rapidly improves with further neurofeedback-training

We demonstrate that lengthening the exposure to visual neurofeedback results in a better control over pathological beta-oscillations, as shown in a stronger and more significant reduction of beta-oscillations by the end of NF 3 (Figure 2). Since one individual achieved a beta-reduction of ∼ 35 % and we did not see any plateau effect in the learning curve by the end of 8 minutes of downregulation training, we hypothesise that stronger beta-reductions could yet be achieved through longer training sessions. Of note, the initial strategy to downregulate (pre-neurofeedback run) on average lead to a higher beta-activity, meaning that controlling deep-brain activity does not seem to be intuitive and that neurofeedback helps in selecting and optimising mental strategies, as all patients reported a different strategy by NF3.

### After training, beta-oscillations can be reduced without neurofeedback

Even in the absence of visual neurofeedback and only after 6 minutes of effective downregulation learning, PD patients still managed to significantly reduce subthalamic beta-activity using their novel neurofeedback-learnt strategies. Performance during this condition (transfer run) was also significantly better than before neurofeedback learning (pre-neurofeedback run) as evaluated by the Wilcoxon’s signed rank test. The fact that the variance increased after turning neurofeedback off could mean that continuous neurofeedback, besides allowing for further learning, also allows for a more efficient control over deep brain activity.

### Motor performance improvements

The analysis of motor performance after employing neurofeedback-learnt strategies revealed promising results in some behavioural outcome metrics. Immediately after downregulation, the pronosupination frequency was significantly higher while movement amplitude was only slightly and non-significantly reduced, still resulting in a larger cumulative angular displacement. Furthermore, the mean torque generated, estimated by the mean absolute angular acceleration and as such a measure for alternating activation of pro- and supinator muscles of the forearm, was significantly increased after downregulation. A similar pattern was observed 2 days later, suggesting that neurofeedback-learnt mental strategies are retained in the long-term and motor performance improvements can be achieved without active neurofeedback.

### Implications for Parkinson’s disease and beyond

We show that self-control over pathological deep brain oscillations can be achieved through DBS-neurofeedback, which in turn improves motor performance. Given the strongly fluctuating nature of PD, neurofeedback-learnt strategies to control pathological deep brain oscillations could help patients cope with situations of transient symptomatic exacerbations: In principle, PD patients could apply neurofeedbacklearnt strategies to overcome Parkinsonian OFF states. Interestingly, the beta-modulatory capacity correlates with disease burden (Fig. 3), possibly reflecting that a pronounced pathological background beta-activity is required for stronger beta-modulations. This finding further motivates the use of deep-brain neurofeedback especially during symptom exacerbations or in severely affected patients. Although our results have to be interpreted cautiously, we provide evidence that endogenous modulation of pathological beta activity may result in improved motor control without the need for dopaminergic treatment or brain stimulation. Thus, neurofeedback could also result in a reduced overall exposure to medication and brain stimulation, thereby combating the development of tolerance as well as potentially having a halting or reversing effect on the natural progression of PD through neuroplasticity by repeated self-activation of neuronal circuits. Along this line, a study conducted in parallel to ours was able to show that neurofeedback of deep brain beta-activity induced changes in the resting oscillatory activity before versus after neurofeedback in the respective direction of neurofeedback ^15^.

These effects can be expected to be more pronounced when longer neurofeedback training is provided using DBS systems that wirelessly transmit signals to an external device ^21^ for real-time neurofeedback. Finally, neurofeedback can reincorporate PD patients into the treatment loop: Compared with current DBS strategies, where PD patients are mere bystanders with little to no control over their burden (and therapy), PD patients could exert neurofeedback-learnt control over pathological deep brain activity in order to reduce stimulation and medication load while simultaneously increasing motor performance through endogenous beta reduction. As DBS has recently also been explored for neurological diseases like epilepsy, obsessive-compulsive disorder, anxiety, depression and attention deficit/hyperactivity disorder, deep brain electrode-guided neurofeedback could be simultaneously studied as a complementing treatment strategy. We showed that not only unidirectional but also bidirectional neurofeedback control can be achieved within a matter of minutes, thus making neurofeedback interesting for cases where a stabilisation or reinforcement of oscillatory patterns is beneficial.

### Outlook

The full extent to which voluntary self-regulation of deep brain oscillatory activity is possible and improves motor outcome yet remains to be unveiled as our study was limited by a short intervention time. We believe that Parkinsonian motor symptoms can be improved by providing longer neurofeedback intervention times, ideally chronic neurofeedback in the context of an implantable device. Improved motor control promoted through these optimised neurofeedback interventions will need to be selectively verified in activities of daily living. Furthermore, future studies should investigate combinatorial approaches and assess whether neurofeedback can reduce the dependence on dopaminergic medication or electrical stimulation, thereby possibly reducing medication side effects or prolonging battery life, respectively.

## Conclusion

This intracranial electrophysiological human study provides the first evidence of patients rapidly gaining real-time control over ongoing deep-brain oscillatory activity through visual neurofeedback. Moreover, neurofeedback-learnt control improves with training duration, is retained in the short and long-term even in the absence of neuro-feedback and improves motor performance. This novel approach could enable PD patients to regain control over aberrant deep brain signalling as well as result in better motor control and, thus, complement current treatment approaches.

## Data Availability

All data referred to in the manuscript are stored locally at the University Hospital Zurich. Data access is strictly restricted as it contains sensitive human subject data.

## Author contributions

Conceptualization: O.B., L.L.I. and R.G.; Methodology: O.B., R.G., C.R.B., and L.L.I; Software: O.B.; Formal Analysis: O.B.; Investigation: O.B., L.S., M.O., and L.L.I; Resources: R.G., L.L.I. and C.R.B; Writing – Original Draft: O.B., L.L.I. and R.G.; Writing – Review & Editing: L.S., M.O., and C.R.B.; Visualization: O.B.; Supervision: L.L.I. and R.G.

## Acknowledgement

This study was supported by grants from the Dr. Wilhelm Hurka Foundation and the Theodor und Ida Herzog Egli Foundation. None of the authors reported any conflict of interest.

